## Supporting Information for "The effect of body image dissatisfaction on goal-directed decision making in a population marked by negative appearance beliefs and disordered eating"

### Decision-making task and questionnaires

#### Exclusion criteria

A range of measures were implemented to exclude participant not taking part in the study in good faith. Firstly, we excluded any participants that took shorter than 1 minute to familiarise themselves with the instruction part of the task. We also excluded any participants, who despite selecting “female” in the pre-screening on Prolific, selected “male”, we allowed three subjects with answers “Other” or “Gender-fluid”. Furthermore, to be included in the study, the participant could only miss a maximum of 10% of trials in each condition. The total reward received at the end of the task had to be higher than two standard deviations subtracted from the average total reward across all participants. Moreover, the average reaction time in each condition had to be higher than two standard deviations from the mean across participants. Lastly, we excluded participants who pressed the same key on more than 95% trials in either stages. At the end of the study, understanding of the structure of the task was tested. We excluded any participants that to the question “If you picked the pirate ship (with skull and cross bones on sails, on the right), which island would you most likely sail to?”, incorrectly answered “blue”. Applying the above criteria amounted to including 67 participants in total, 32 in the HC and 35 in the ED group.

#### Body types

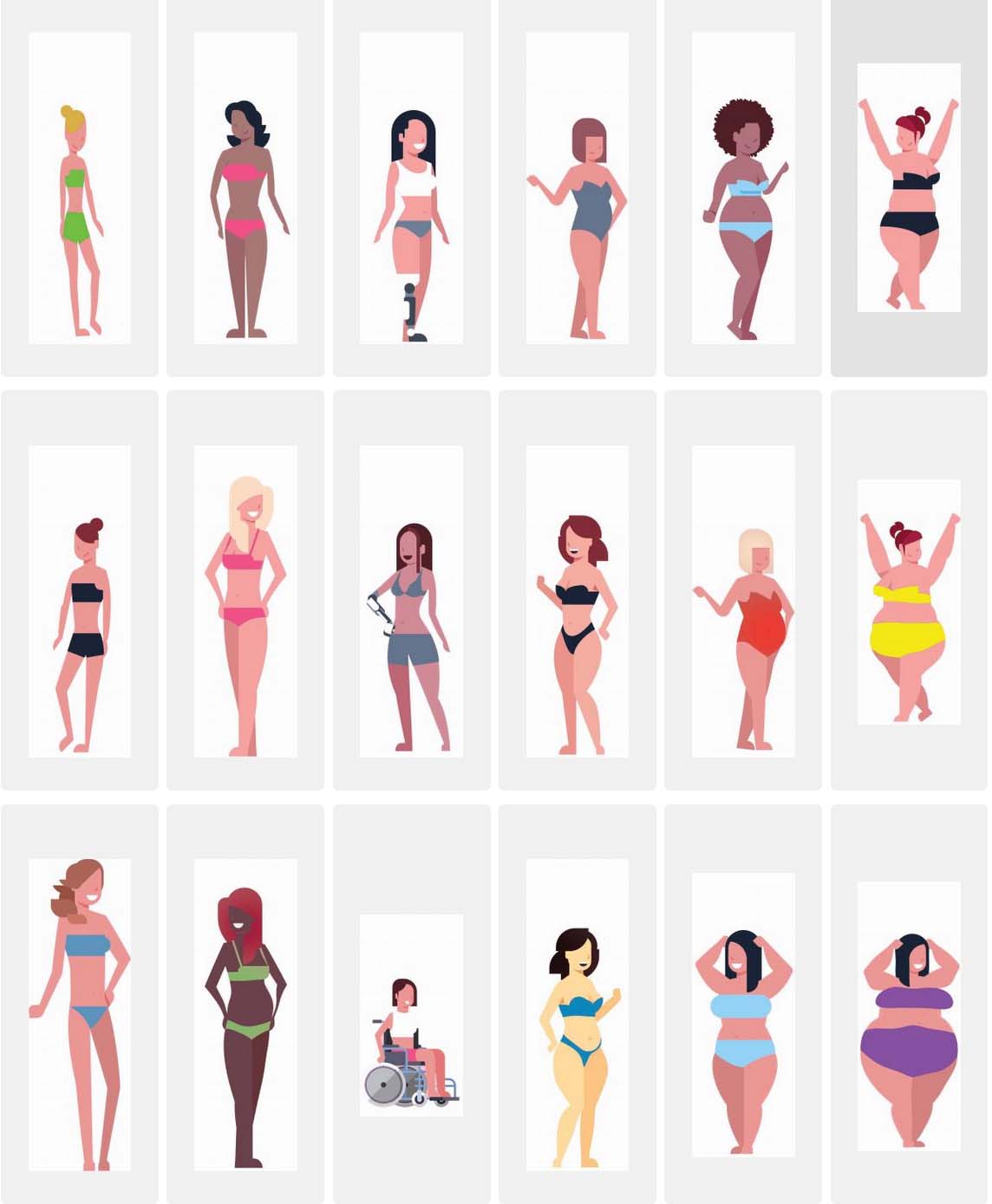

**S1 Fig.** 18 different types of body that participants could select as most similar to their own.

### Computational modelling

#### Parameter recovery

To ensure reliable parameter estimates, it is advisable to perform parameter recovery to test if the range of parameters is well recovered (1). In parameter recovery, we first simulate some fake data using our model and a set of parameters drawn from a distribution. Then, we try to recover the parameters that generated the fake data using our model fitting technique. To see how reliable the modelling process is, we compare the original parameters ($X$) with the recovered $Y$ ones by calculating the Pearson correlation coefficient (PCC), $r$: $r=\frac{cov\left( X,Y \right)}{\sigma_{X},\sigma_{Y}}$, where $cov(X,Y)$ is the covariance between two sets of parameters and $\sigma$ is the standard deviation. The higher $r$, the better and more reliable our methods are, and can be categorised as: poor (if $r$ <0.5); fair (if 0.5< $r$ <0.75); good (0.75< $r$ <0.9); excellent (if $r$ >0.9) (2). We did not perform parameter recovery prior to data collection as it can be affected by the parameter range obtained (1). However, previous studies show the recovery to range from fair to excellent (3,4). As such, we focused on the parameter ranges estimated from the data to obtain a more accurate picture of the parameter recovery analysis.

#### Gaussian random walk

The reward probabilities evolved according to a Gaussian random walk. In the first condition the participants were faced with, the reward probabilities were initialised in a range (for each chest from left to right: $p_{11},p_{12}, p_{21},p_{22}$):

$$\left[ 0.58,0.72 \right],\left[ 0.31,0.45 \right],\left[ 0.31,0.45 \right],\left[ 0.58,0.72 \right].$$

In the second condition, the initialisation of reward probabilities was:

$$[0.31,0.45],[0.58,0.72],[0.58,0.72],[0.31,0.45].$$

The practice stage was initialised differently as:

$$[0.31,0.45],[0.58,0.72],[0.31,0.45],[0.58,0.72].$$

The general formula of a Gaussian random walk takes a form:

$$p^{\left( t+1 \right)}=p^{(t)}+\sigma\times\mathcal{N}\left( 0,1 \right),$$

where$p^{(t)}$ is a reward probability at a current trial $t$, $\sigma$ is the standard deviation of the random walk, $\mathcal{N}\left( 0,1 \right)$ is a random variable from a standard normal distribution.

To make sure the reward probabilities always stay within a $(0.25,0.75)$range, any resulting value that was above or below the range was transformed:

$$p^{(t)}=2\times r_{hi/lo}-p^{\left( t \right)},$$

where $r_{hi/lo}$ $0.25$ or $0.75$.

### Supplementary results

#### Parameter recovery

We carried out parameter recovery of for the model, for each group and condition 10 times (using 1000 samples for each of four chains). The average PCCs, $r,$from 10 runs (each with a different random seed) of the procedure are presented in S1 Table.

**S1 Table. Parameter recovery from collected data.**

|  | **Parameters** | | | | |
| --- | --- | --- | --- | --- | --- |
| mean $\pm$ SD | $\beta_{\mathrm{MB}}$ | $\beta_{\mathrm{MF}}$ | $\beta_{2}$ | $\alpha$ | $\rho$ |
| $r$ (ED,NT) | 0.53 ± 0.13 | 0.83 ± 0.07 | 0.93 ± 0.03 | 0.94 ± 0.02 | 0.90 ± 0.04 |
| $r$ (ED,BID) | 0.51 ± 0.09 | 0.81 ± 0.05 | 0.92 ± 0.04 | 0.95 ± 0.02 | 0.91 ± 0.04 |
| $r$ (HC,NT) | 0.58 ± 0.12 | 0.82 ± 0.05 | 0.90 ± 0.03 | 0.98 ± 0.01 | 0.87 ± 0.05 |
| $r$ (HC,BID) | 0.72 ± 0.05 | 0.81 ± 0.08 | 0.92 ± 0.02 | 0.92 ± 0.02 | 0.92 ± 0.04 |

Mean and standard deviation of PCC, $r,$ of each model parameter, in each group and condition after data collection

#### Task performance

**S2 Table. Summary of model-independent task performance measures for two groups.**

| **Measures** | **HC (n=36)**  **Mean** $\boldsymbol{\pm}$**SD** | **ED (n=37)**  **Mean** $\boldsymbol{\pm}$**SD** | **t value** | **p value** |
| --- | --- | --- | --- | --- |
| reward: NT | 81.94 ± 9.79 | 77.43 ± 8.99 | 1.97 | 0.0536 |
| reward: BID | 76.59 ± 8.08 | 78.63 ± 9.07 | -0.97 | 0.3377 |
| total reward | 158.53 ± 12.63 | 156.06 ± 13.08 | 0.79 | 0.4348 |
| RT1: all | 0.28 ± 0.07 | 0.31 ± 0.07 | -1.31 | 0.1932 |
| RT2: all | 0.39 ± 0.09 | 0.42 ± 0.13 | -1.12 | 0.2683 |
| RT: all | 0.34 ± 0.07 | 0.36 ± 0.09 | -1.30 | 0.1983 |
| RT1: neutral | 0.29 ± 0.08 | 0.3 ± 0.08 | -0.71 | 0.4776 |
| RT2: neutral | 0.39 ± 0.11 | 0.42 ± 0.15 | -1.00 | 0.3201 |
| RT: neutral | 0.34 ± 0.09 | 0.36 ± 0.11 | -0.96 | 0.3420 |
| RT1: BID | 0.28 ± 0.08 | 0.31 ± 0.09 | -1.60 | 0.1134 |
| RT2: BID | 0.39 ± 0.1 | 0.42 ± 0.13 | -1.02 | 0.3110 |
| RT: BID | 0.33 ± 0.08 | 0.37 ± 0.1 | -1.39 | 0.1694 |

Average total reward during the neutral and BID conditions, average total reward after a whole task, mean reaction time in the first and second stage and overall (RT1, RT2, RT) in the neutral, BID, and across both conditions. T- and p-values of the two-sample t-tests for difference in performance between groups are included.

#### Model parameters

**S3 Table. Mixed effects linear regression analysis of model-based learning parameter** $\boldsymbol{\rho}$

| **Effects** | **Estimate** | **SE** | **t value** | **p value** |
| --- | --- | --- | --- | --- |
| Intercept (HC, NT condition) | 1.41 | 0.17 | 8.30 | **<0.001*** |
| ED group | -0.19 | 0.24 | -0.79 | 0.4320 |
| BID condition | 0.00 | 0.11 | -0.02 | 0.9830 |
| Age | 0.13 | 0.12 | 1.14 | 0.2590 |
| ED group $\times$ BID condition | -0.03 | 0.16 | -0.22 | 0.8270 |

Group, condition, age are treated as fixed-effect covariates per subject.

#### Simulated raw choice data analysis

##### Mixed-effects logistic regression analysis – simulated data.

To check whether the RL model is able to capture the contribution of model-based and model-free system in the recruited populations, ensuring meaningful and interpretable results, we used the individually estimated parameters to simulate fake choice data (5), which was then analysed used mixed-effects logistic regression, looking for the effects of reward and its interaction with transition. This essentially mimics the experiment, where the RL agent take on the role of the participant. In the model with the interaction between group and condition as a variable (S4 Table), we detected model-free and model-based contribution in the neutral condition in HC (p-value=0.013, p-value<0.001, respectively). Moreover, the model also detected reduced model-based contribution in the ED group as compared to HC group in the neutral condition (p-value=0.013). There were no differences between conditions in either group. The estimated probabilities (frequency-based) for each case in each group and condition are depicted in S2 Fig.

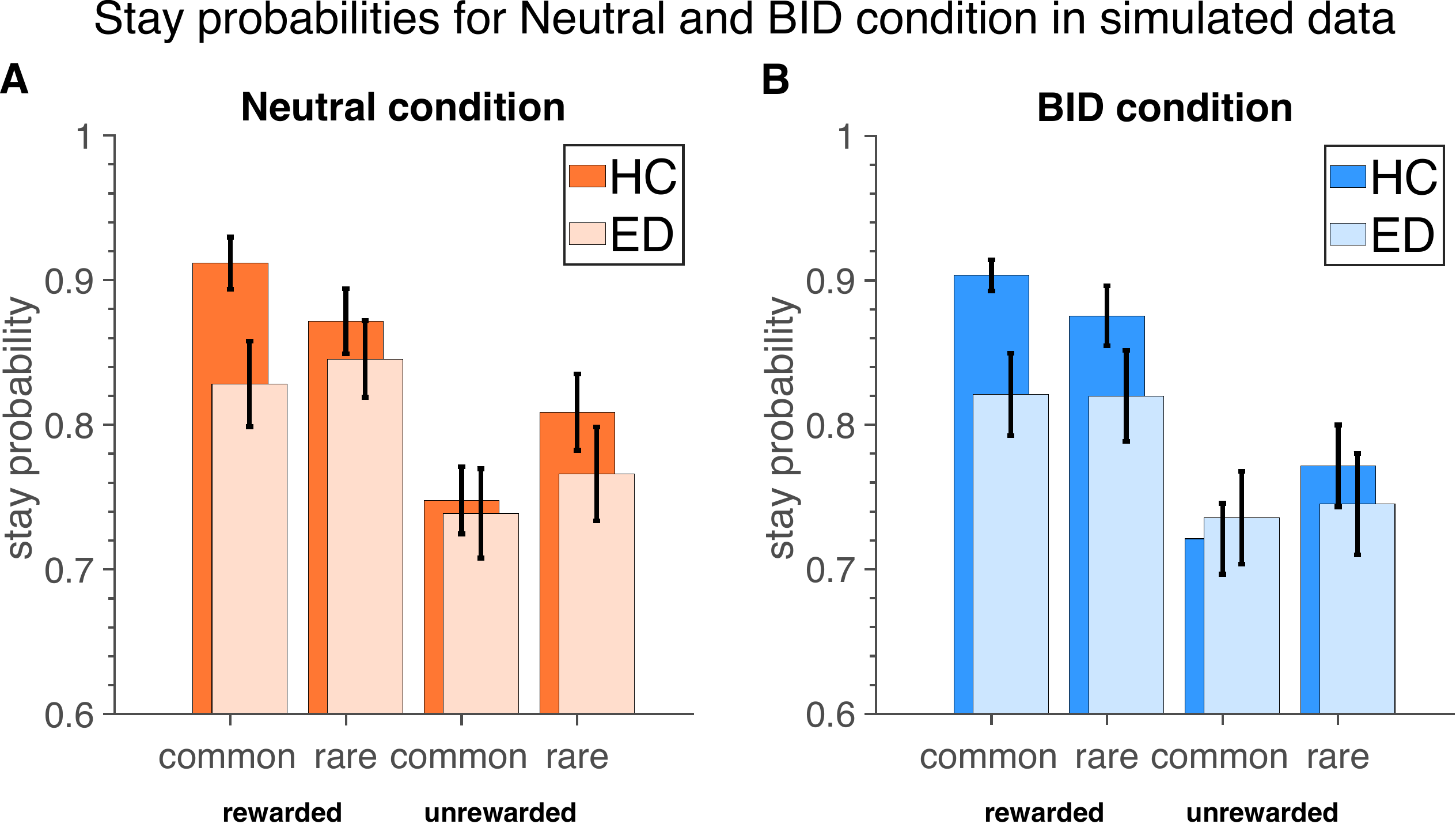

**S2 Fig. Stay probabilities in the simulated data.** (A) Neutral condition. (B) BID condition.

**S4 Table. Random effects logistic regression for probability of staying (simulated data).**

| **Effects** | **Estimate** | **SE** | **z value** | **p value** |
| --- | --- | --- | --- | --- |
| Intercept (HC, conditionNT) | 1.64 | 0.20 | 8.16 | **<0.001*** |
| reward | 0.52 | 0.21 | 2.48 | **0.013*** |
| transition | -0.42 | 0.13 | -3.20 | **0.001*** |
| groupED | -0.36 | 0.29 | -1.23 | 0.217 |
| conditionBID | -0.27 | 0.14 | -2.00 | **0.045*** |
| age_z | 0.09 | 0.14 | 0.68 | 0.496 |
| reward × transition | 0.82 | 0.23 | 3.63 | **<0.001*** |
| groupED × conditionBID | 0.14 | 0.19 | 0.76 | 0.446 |
| reward × groupED | 0.06 | 0.29 | 0.22 | 0.826 |
| reward × conditionBID | 0.26 | 0.21 | 1.25 | 0.213 |
| reward × age_z | 0.01 | 0.13 | 0.12 | 0.907 |
| transition × groupED | 0.24 | 0.18 | 1.31 | 0.190 |
| transition × conditionBID | 0.12 | 0.16 | 0.77 | 0.444 |
| transition × age_z | 0.00 | 0.07 | -0.05 | 0.961 |
| reward × groupED × conditionBID | -0.35 | 0.29 | -1.23 | 0.220 |
| transition × groupED × conditionBID | -0.02 | 0.22 | -0.09 | 0.929 |
| reward × transition × groupED | -0.77 | 0.31 | -2.49 | **0.013*** |
| reward × transition × conditionBID | -0.22 | 0.25 | -0.87 | 0.382 |
| reward × transition × age_z | -0.06 | 0.13 | -0.44 | 0.660 |
| reward × transition × groupED × conditionBID | 0.26 | 0.34 | 0.77 | 0.443 |

*Note*. Group, condition, age are treated as fixed-effect covariates per subject.

##### Frequency based-probability regression – simulated data.

Furthermore, comparing the simulated probabilities of staying after common and rewarded trials using the frequency calculation in random effects linear regression (S5 Table), we again found a weaker model-based learning capacity in ED (p-value = 0.033). There were no differences between conditions in either group.

**S5 Table. Random effects linear regression for probability of staying (frequency-based; simulated data).**

| **Effects** | **Estimate** | **SE** | **t value** | **p value** |
| --- | --- | --- | --- | --- |
| Intercept (HC, neutral) | 0.91 | 0.03 | 35.61 | **<0.001*** |
| groupED | -0.08 | 0.04 | -2.17 | **0.033*** |
| conditionBID | -0.01 | 0.02 | -0.46 | 0.649 |
| age_z | 0.00 | 0.02 | -0.25 | 0.803 |
| groupED:conditionBID | 0.00 | 0.03 | 0.04 | 0.968 |

*Note*. Age is treated as fixed-effect covariates per subject.

#### MB score analysis – simulated raw choice data

Lastly, we calculated MB scores in the simulated data (S3 Fig). These were regressed against group and condition variables (S6 Table). The simulated data captured reduced MB scores in the ED group in the neutral condition (p-value = 0.040).

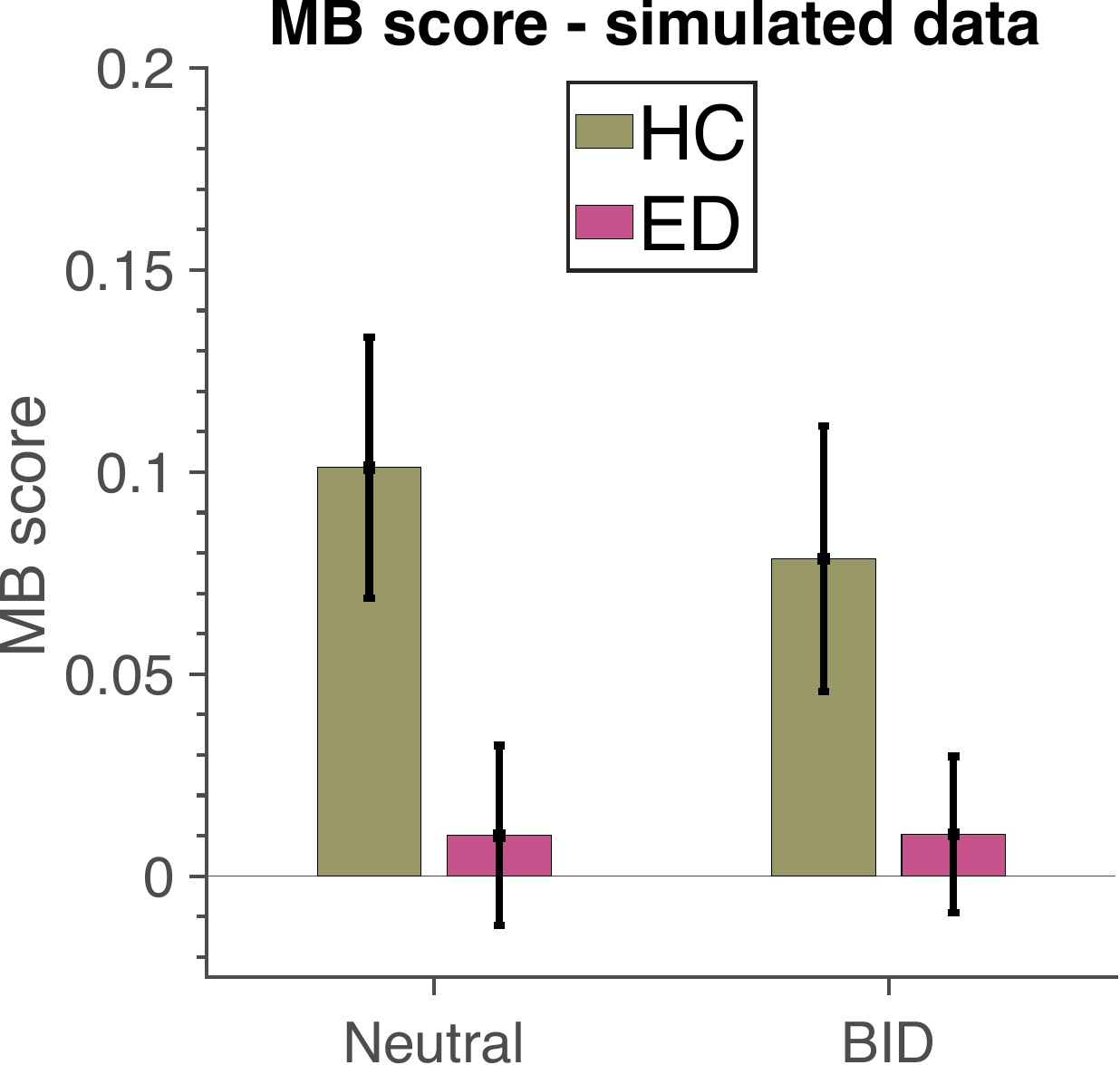

**S3 Fig. MB scores** per group and condition. (A) collected and (B) simulated data.

**S6 Table. Random effects linear regression for MB score (simulated data).**

| **Effects** | **Estimate** | **SE** | **t value** | **p value** |
| --- | --- | --- | --- | --- |
| Intercept (HC, conditionNT) | 0.10 | 0.03 | 3.42 | **<0.001*** |
| groupED | -0.09 | 0.04 | -2.08 | **0.040*** |
| conditionBID | -0.02 | 0.03 | -0.73 | 0.471 |
| age_z | -0.01 | 0.02 | -0.39 | 0.699 |
| groupED:conditionBID | 0.02 | 0.04 | 0.53 | 0.598 |

#### Correlation between questionnaire scores

**S7 Table. Correlation between questionnaire scores**

| **Covariates** | | | |
| --- | --- | --- | --- |
|  | **EAT-26** | **AAI** | **OCI-R** |
| **EAT-26** | 1 | 0.896 (p-value<**0.001***) | 0.768 (p-value<**0.001***) |
| **AAI** |  | 1 | 0.814 (p-value<**0.001***) |
| **OCI-R** |  |  | 1 |

Correlation coefficients r and P-values for the hypothesis of no relationship between the covariates (EAT-26, AAI, OCI-R score all z-scored) are included.

### References

1.	Wilson RC, Collins AG. Ten simple rules for the computational modeling of behavioral data. eLife. 2019;8:e49547.

2.	White CN, Servant M, Logan GD. Testing the validity of conflict drift-diffusion models for use in estimating cognitive processes: A parameter-recovery study. Psychon Bull Rev. 2018;25(1):286–301.

3.	Shahar N, Hauser TU, Moutoussis M, Moran R, Keramati M, NSPN consortium, et al. Improving the reliability of model-based decision-making estimates in the two-stage decision task with reaction-times and drift-diffusion modeling. Gershman SJ, editor. PLoS Comput Biol. 2019;15(2):e1006803.

4.	Ballard IC, McClure SM. Joint modeling of reaction times and choice improves parameter identifiability in reinforcement learning models. Journal of Neuroscience Methods. 2019;317:37–44.

5.	Foerde K, Daw ND, Rufin T, Walsh BT, Shohamy D, Steinglass JE. Deficient Goal-Directed Control in a Population Characterized by Extreme Goal Pursuit. Journal of Cognitive Neuroscience. 2020;1–19.
